## Supplementary Material for "WASH interventions and child diarrhea at the interface of climate and socioeconomic position in Bangladesh"

**Supplementary Table 1. Summary of household characteristic means and standard deviation by wealth index quintiles.** The household asset-based characteristics were included in the construction of the wealth index through a principal component analysis. The factor loading was the eigenvector from the first principal component.

| Asset-based characteristics included in the wealth index | Wealth Index Quintiles | | | | | Factor Loading |
|---|---|---|---|---|---|---|
| | 1 (N=1110) | 2 (N=1110) | 3 (N=1110) | 4 (N=1109) | 5 (N=1112) | |
| | Mean (SD) | Mean (SD) | Mean (SD) | Mean (SD) | Mean (SD) | |
| **Land owned in acres (Mean, SD)** | 0.07 (0.07) | 0.10 (0.10) | 0.13 (0.17) | 0.17 (0.33) | 0.26 (0.34) | 0.09 |
| **Improved wall material (wood, brick, thin)** | 0.64 (0.48) | 0.66 (0.48) | 0.79 (0.41) | 0.77 (0.42) | 0.74 (0.44) | 0.06 |
| **Improved floor material (wood, concrete)** | 0.003 (0.052) | 0.01 (0.11) | 0.05 (0.21) | 0.09 (0.28) | 0.38 (0.49) | 0.17 |
| **Household has electricity** | 0.14 (0.35) | 0.35 (0.48) | 0.52 (0.50) | 0.93 (0.25) | 1.0 (0.07) | 0.42 |
| **Household has refrigerator** | 0.00 (0.00) | 0.001 (0.030) | 0.003 (0.052) | 0.03 (0.17) | 0.36 (0.48) | 0.15 |
| **Household has bicycle** | 0.08 (0.27) | 0.22 (0.41) | 0.34 (0.47) | 0.40 (0.49) | 0.51 (0.50) | 0.21 |
| **Household has motorcycle** | 0.002 (0.042) | 0.004 (0.060) | 0.03 (0.16) | 0.06 (0.24) | 0.23 (0.42) | 0.11 |
| **Household has sewing machine** | 0.01 (0.09) | 0.03 (0.18) | 0.05 (0.21) | 0.08 (0.27) | 0.16 (0.36) | 0.07 |
| **Has black and white or colored TV** | 0.002 (0.042) | 0.04 (0.20) | 0.12 (0.33) | 0.39 (0.49) | 0.94 (0.25) | 0.41 |
| **Has one or more wardrobe** | 0.01 (0.09) | 0.05 (0.22) | 0.09 (0.28) | 0.16 (0.37) | 0.52 (0.50) | 0.22 |
| **Has one or more table** | 0.22 (0.42) | 0.66 (0.48) | 0.85 (0.36) | 0.92 (0.28) | 0.98 (0.14) | 0.34 |
| **Has one or more chair** | 0.19 (0.39) | 0.65 (0.48) | 0.87 (0.34) | 0.94 (0.24) | 1.0 (0.07) | 0.36 |
| **Has one or more khat (type of bed)** | 0.07 (0.25) | 0.37 (0.48) | 0.71 (0.46) | 0.91 (0.29) | 1.0 (0.07) | 0.44 |
| **Has one or more chouki (type of chair)** | 0.95 (0.22) | 0.83 (0.38) | 0.77 (0.42) | 0.77 (0.42) | 0.63 (0.48) | -0.14 |
| **Has one or more mobile** | 0.56 (0.50) | 0.82 (0.38) | 0.93 (0.26) | 0.98 (0.16) | 0.99 (0.09) | 0.19 |

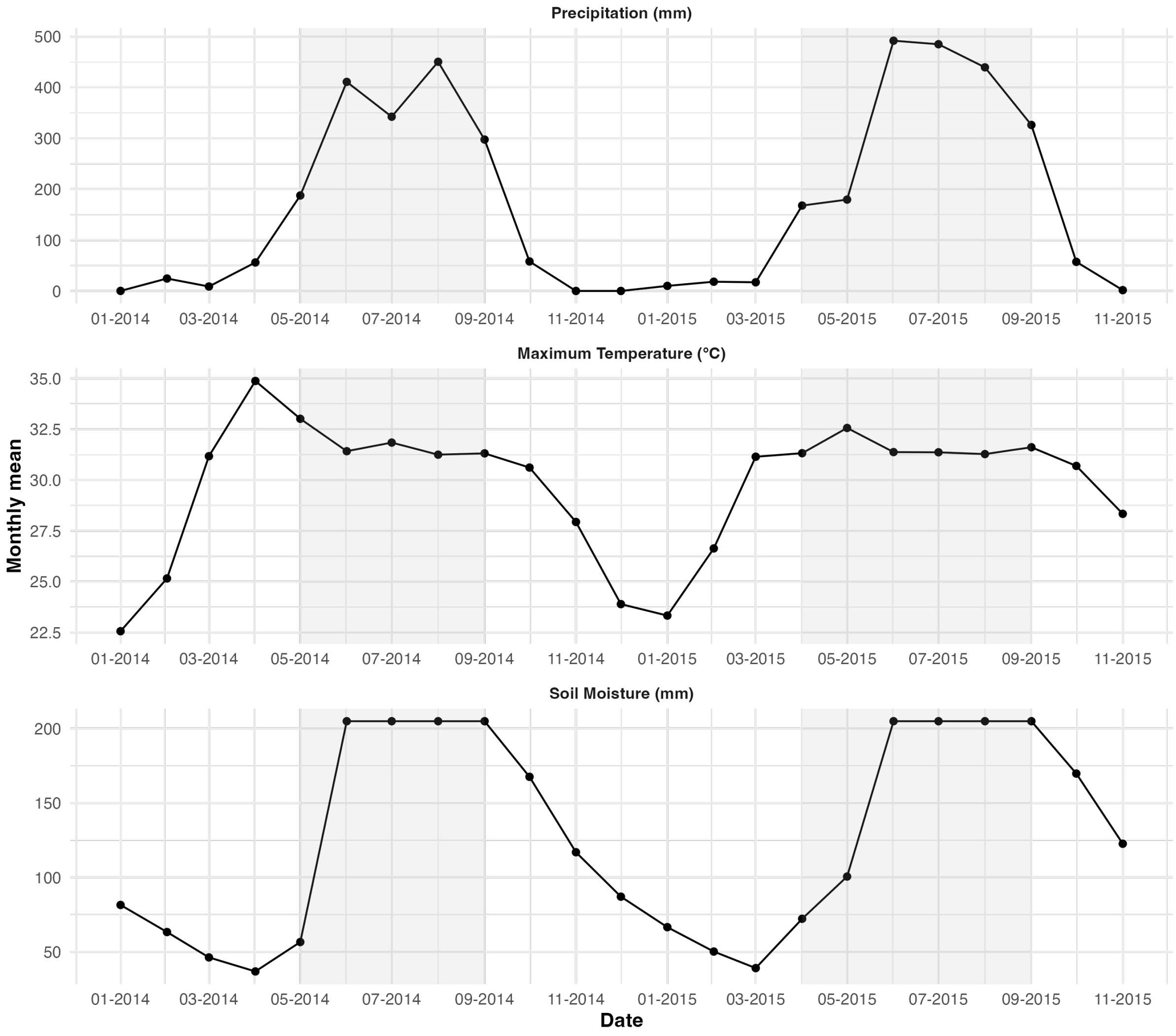


**Supplementary Fig. 1: Climate characteristics during the study.** Monthly mean by precipitation, maximum temperature and soil moisture during the study trial. Shaded areas illustrate the monsoon season defined as weeks with elevated precipitation (May 27 – September 27, 2014, and April 1 – September 26, 2015).

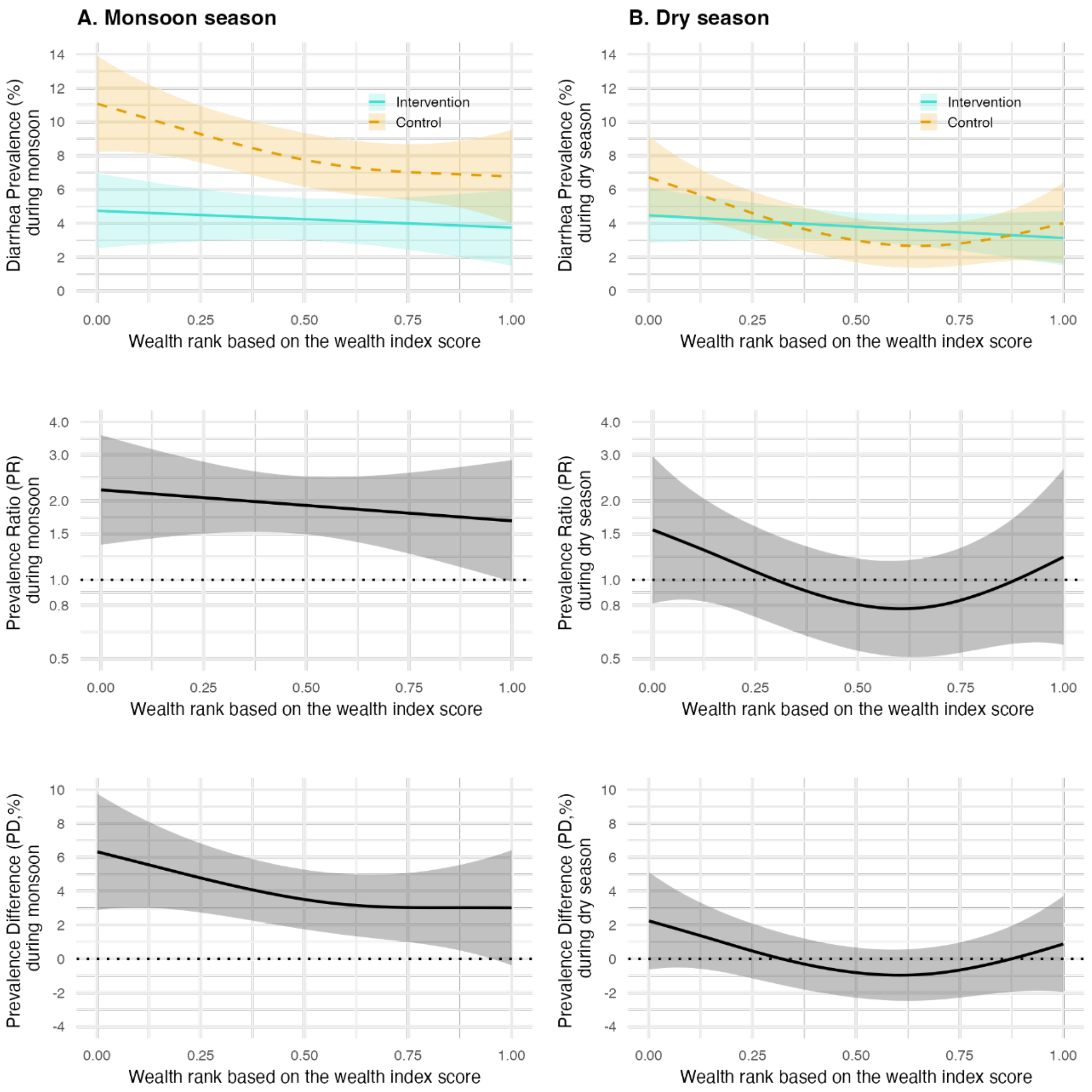


**Supplementary Fig. 2: Effect of WASH interventions by socioeconomic position using a continuous wealth score.** Left panels show estimates during the monsoon season. Right panels show estimates during the dry season. The Y-axes for the prevalence ratios are on a log scale. Shaded areas represent 95% confidence intervals.

**Supplementary Table 2. Summary of baseline characteristics by maternal education.**

| Baseline characteristics | Maternal Education | | |
|---|---|---|---|
| | No education (N=259) | Primary (1-5y) (N=591) | Secondary (>5y) (N=758) |
| **Asset-based characteristics included in the wealth index** | | | |
| **Land owned in acres (Mean, SD)** | 0.0723 (0.0742) | 0.0946 (0.118) | 0.160 (0.212) |
| **Improved wall material (wood, brick, thin)** | 172 (66.4%) | 468 (79.2%) | 541 (71.4%) |
| **Improved floor material (wood, concrete)** | 4 (1.5%) | 31 (5.2%) | 106 (14.0%) |
| **Household has electricity** | 99 (38.2%) | 287 (48.6%) | 509 (67.2%) |
| **Household has refrigerator** | 0 (0%) | 8 (1.4%) | 100 (13.2%) |
| **Household has bicycle** | 52 (20.1%) | 109 (18.4%) | 279 (36.8%) |
| **Household has motorcycle** | 0 (0%) | 18 (3.0%) | 81 (10.7%) |
| **Household has sewing machine** | 0 (0%) | 11 (1.9%) | 75 (9.9%) |
| **Has black and white or colored TV** | 15 (5.8%) | 98 (16.6%) | 268 (35.4%) |
| **Has one or more wardrobe** | 12 (4.6%) | 48 (8.1%) | 171 (22.6%) |
| **Has one or more table** | 140 (54.1%) | 394 (66.7%) | 600 (79.2%) |
| **Has one or more chair** | 139 (53.7%) | 372 (62.9%) | 625 (82.5%) |
| **Has one or more khat (type of bed)** | 89 (34.4%) | 309 (52.3%) | 556 (73.4%) |
| **Has one or more chouki (type of chair)** | 228 (88.0%) | 455 (77.0%) | 568 (74.9%) |
| **Has one or more mobile** | 179 (69.1%) | 479 (81.0%) | 680 (89.7%) |
| **Asset-based characteristics not included in the wealth index** | | | |
| **Primary water source: shallow tubewell** | 179 (69.1%) | 447 (75.6%) | 557 (73.5%) |
| **Store drinking water** | 136 (52.5%) | 295 (49.9%) | 331 (43.7%) |
| **Reported treating water today/tomorrow** | 0 (0%) | 2 (0.3%) | 2 (0.3%) |
| **Own their latrine** | 91 (35.1%) | 214 (36.2%) | 371 (48.9%) |
| **Latrine has concrete slab** | 203 (78.4%) | 504 (85.3%) | 708 (93.4%) |
| **Latrine has functional water seal** | 31 (12.0%) | 81 (13.7%) | 212 (28.0%) |
| **No visible feces on floor of latrine** | 88 (34.0%) | 193 (32.7%) | 361 (47.6%) |
| **Has a potty for defecation** | 3 (1.2%) | 26 (4.4%) | 85 (11.2%) |
| **Primary handwashing location has water/soap** | 19 (7.3%) | 81 (13.7%) | 209 (27.6%) |
| **Household has radio** | 6 (2.3%) | 13 (2.2%) | 44 (5.8%) |
| **Has one or more clock** | 39 (15.1%) | 133 (22.5%) | 352 (46.4%) |

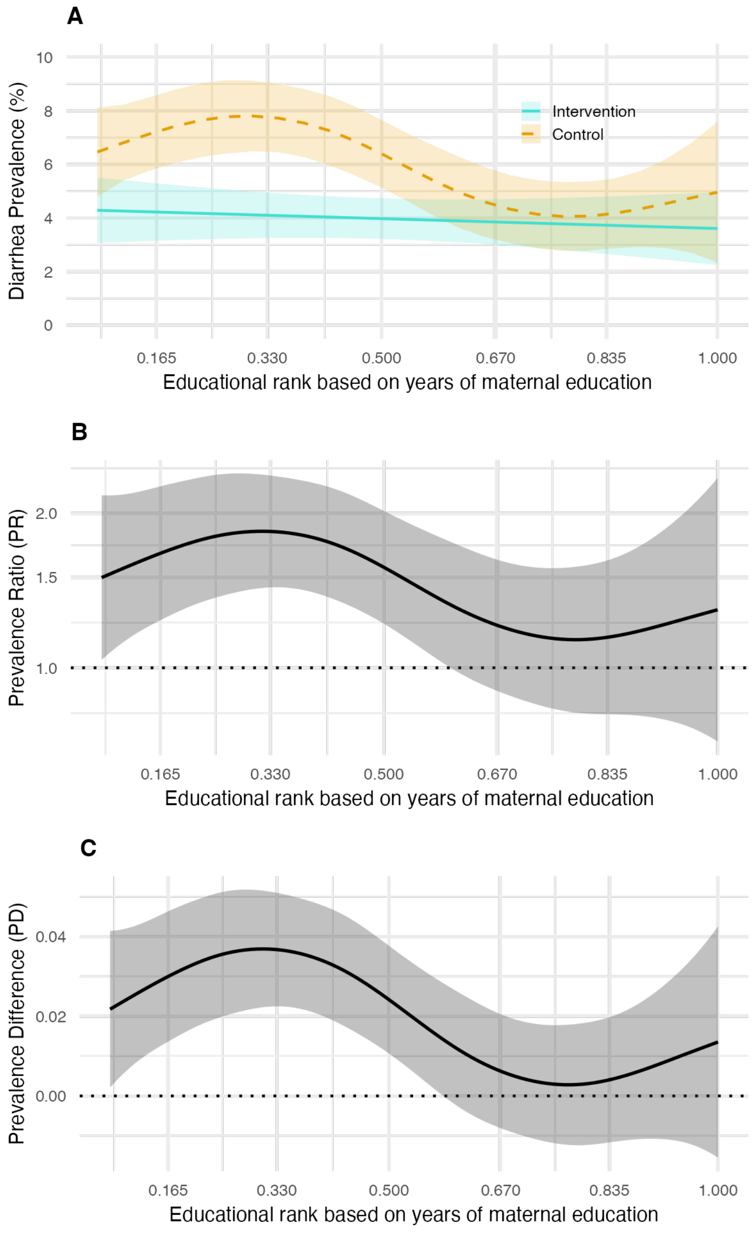


**Supplementary Fig. 3: Effect of WASH interventions on diarrhea by mother's educational rank based on the number of years of education. A:** Diarrhea prevalence along the continuous educational rank in the control and intervention groups. **B:** Prevalence ratio of child diarrhea along the continuous educational rank in the control and intervention groups. **C:** Prevalence difference of child diarrhea along the continuous educational rank in the control and intervention groups. Shaded areas represent 95% confidence intervals.

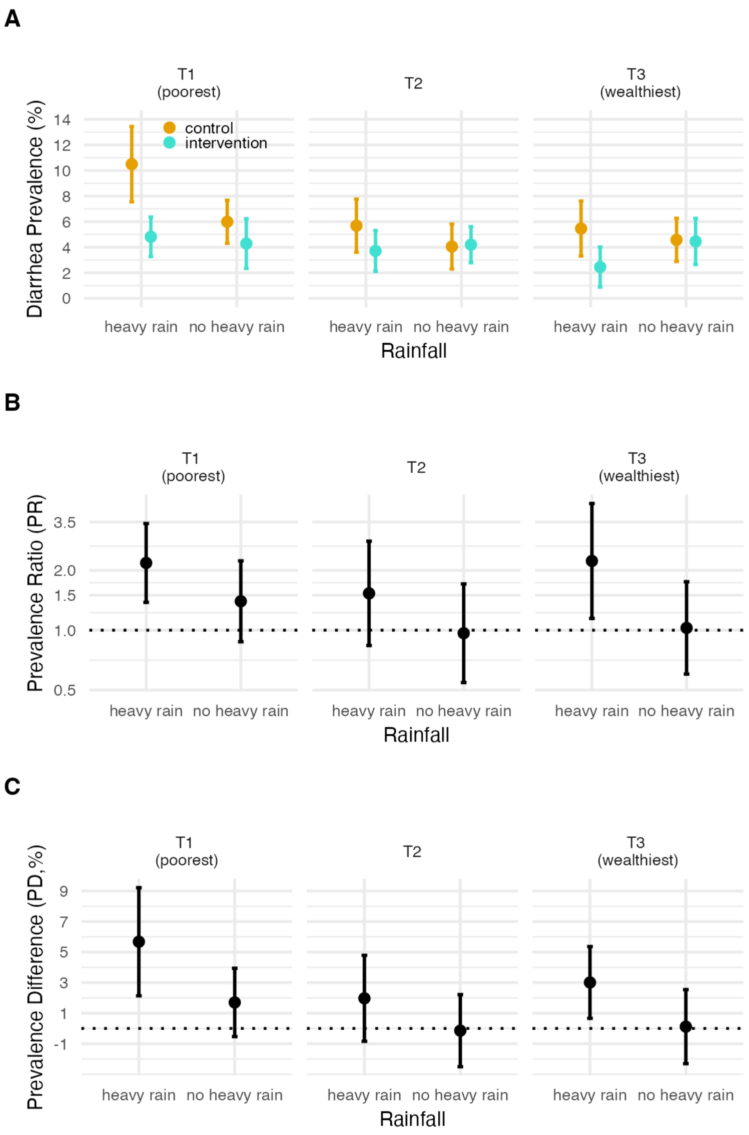


**Supplementary Fig. 4: Effect of WASH interventions by socioeconomic position and rainfall. A:** Diarrhea prevalence along the tertiles of wealth index in the event of heavy rain versus no heavy rain in the control and intervention groups. **B:** Prevalence ratio of child diarrhea along the tertiles of wealth index in the event of heavy rain versus no heavy rain in the control and intervention groups. Y-axis is on a log scale**. C:** Prevalence difference of child diarrhea along the tertiles of wealth index in the event of heavy rain versus no heavy rain in the control and intervention groups. Error bars represent 95% confidence intervals.

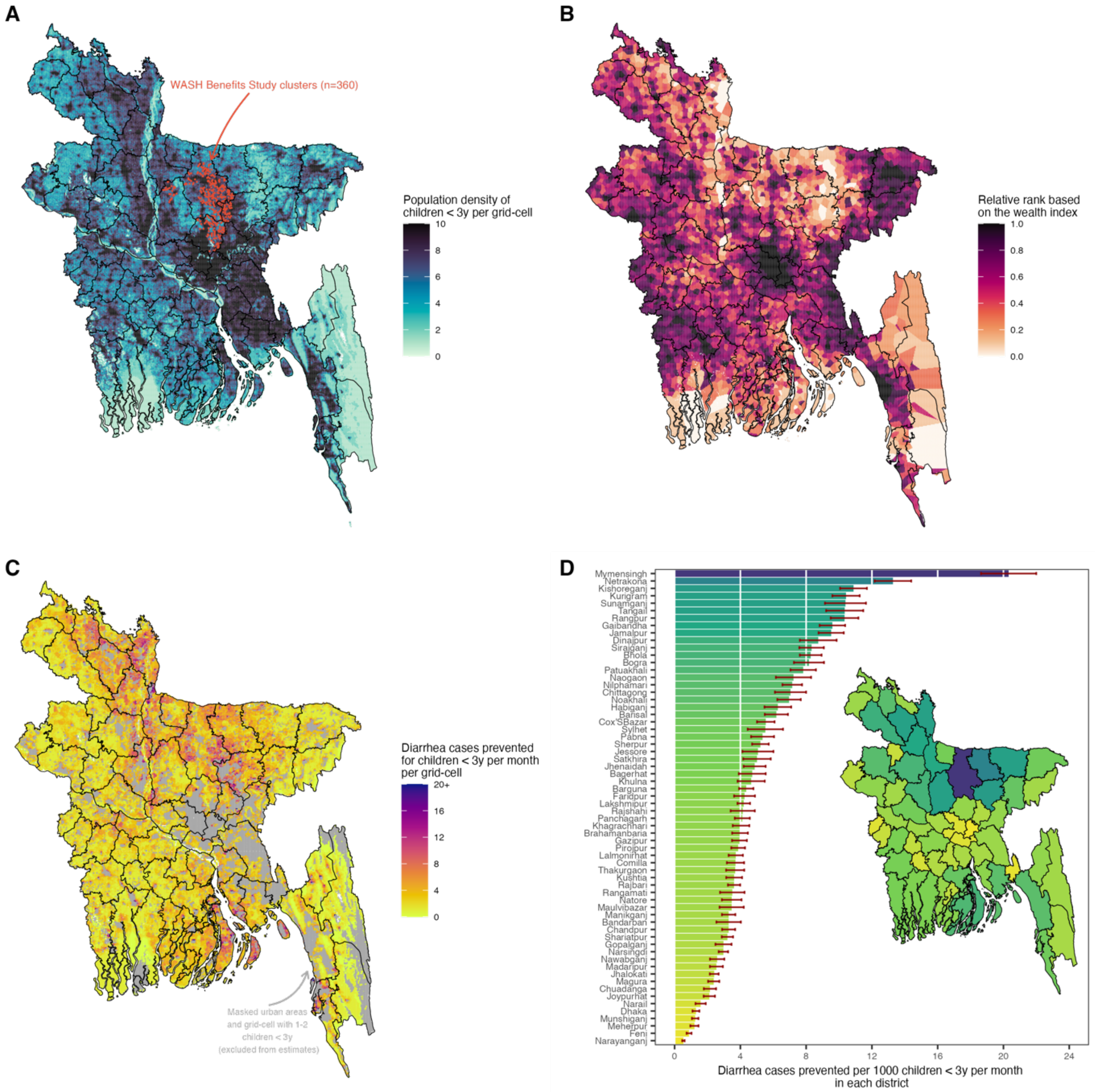


**Supplementary Fig. 5: Diarrhea cases prevented by a combined WASH intervention along a continuous wealth gradient per grid-cell averaged over the entire year at a 1-kilometer resolution and per district in rural Bangladesh. A:** WASH Benefits Study clusters that were included in the analysis (n=360) and gridded population density of children <3 years estimated from WorldPop. **B:** Distribution of relative wealth rank based on the Demographic and Health Surveys wealth index with Bayesian model-based geostatistics and high resolution gridded spatial covariates and aggregated phone data per grid-cell at 1-km resolution estimated from WorldPop. **C:** The projected diarrhea cases prevented by combined WASH per month by wealth per grid-cell at 1-km resolution. We masked non-urban areas based on the Global Human Settlement and grid-cell with only 1-2 children < 3 years. **D:** The projected diarrhea cases prevented by combined WASH per 1000 children under 3 years per month in panel C aggregated for each administrative district. Error bars represent 95% confidence intervals.

**Supplementary Text 1.** We assessed the effect modification of WASH by socioeconomic position, monsoon season and jointly by socioeconomic position and monsoon season. We assessed the effect modification by comparing the models with and without the interaction term through a Wald-type F test to test for statistical significance.

### Effect modification by socioeconomic position

$$P(diarrhea)\sim f(WASH, SEP\colon WASH)$$

*versus*

$$P(diarrhea)\sim f(WASH, SEP)$$

### Effect modification by monsoon season

$$P(diarrhea)\sim f(WASH, monsoon\ season\colon WASH)$$

*versus*

$$P(diarrhea)\sim f(WASH, monsoon\ season)$$

### Joint effect modification by socioeconomic position and monsoon season

$$P(diarrhea)\sim f(WASH, SEP, monsoon\ season, SEP\colon WASH, monsoon\ season\colon WASH,$$
$$SEP\colon monsoon\ season\colon WASH)$$

*versus*

$$P(diarrhea)\sim f(WASH, SEP, monsoon\ season),$$

where SEP = socioeconomic position, WASH = Water, Sanitation and Handwashing.
